# Unintended events in long-distance neonatal interhospital transport in Western Australia: A comparison of neonatal specialist and non-neonatal specialist transport teams

**DOI:** 10.1101/2023.09.26.23296136

**Authors:** Jacqueline Gardiner, Kylie McDonald, Joanne Blacker, Sam Athikarisamy, Mary Sharp, Jonathan Davis

**Affiliations:** Newborn Emergency Transport Service WA, Perth Children’s Hospital, Child and Adolescent Health Service, Perth, Western Australia; Neonatal Directorate, Perth Children’s Hospital, Child and Adolescent Health Service, Perth, Western Australia; Neonatal Directorate, King Edward Memorial Hospital, Child and Adolescent Health Service, Perth, Western Australia; University of Western Australia, Perth, Western Australia

**Keywords:** Outborn, Neonatal Retrieval, Safety, Aeromedical, Adverse events, Thermoregulation, Neonatal nursing

## Abstract

**Objective:** To compare unintended events in transport performed by neonatal specialist (NS) and non-neonatal specialist (NNS) teams in Western Australia (WA).

Study design: Retrospective comparison of neonatal transports from the Kimberley and Pilbara regions (WA) to tertiary services, King Edward Memorial (perinatal) and Perth Children’s Hospital NICU, in Perth (state capital, distance up to 2200km) between January 2018 - June 2021. NS teams travelled from the tertiary to the referring center and back. NNS travelled from the referring to the tertiary center. Transport time, team composition, total unintended clinical, endotracheal tube (ETT), and logistic events were compared. Categorial data are expressed as numbers (%) and compared by chi-squared test; continuous data are expressed as median (interquartile range) and compared by Mann-Whitney U test.

**Results:** During the study period, 3,709 infants were transported in WA to tertiary services for escalation of care: 119(3.2%) from the Kimberley and Pilbara, 49 with NS and 70 with NNS teams. NNS had shorter transport times than NS teams (508(435 – 609) vs 871(615 – 735) min; p<0.00001). Neonatal nurses were on NS more than NNS teams (36/49(73.5%) vs 6/70(9%); p<0.001). Total unintended clinical and ETT events were less in NS vs. NSS teams (28/49(57%) vs. 54/70(77%); p=0.02) and (0/26(0%) vs. 7/23(30%); p=0.004). Unintended logistic events were greater involving NS than NNS teams (31/49 (63%) vs. 33/70 (47%); p=0.05).

**Conclusion:** Although NS teams had longer transport times and more unintended logistic events, there were fewer unintended clinical and ETT events. NS teams should be considered as the first choice to undertake neonatal transport.

## Introduction

Emergency transport is a critical component of centralized neonatal care. Centralization of preterm and low birth weight infants has been recommended worldwide for many years (1), improving outcome survival and morbidity. (2-6). Despite best efforts to transfer high-risk women antenatally, some infants require stabilization and transfer to a tertiary-level Neonatal Intensive Care Unit (NICU) after birth. Infants cared for by healthcare staff inexperienced at newborn resuscitation take longer to intubate, have more attempts to establish intravenous access and have a higher rate of hypothermia (7).

Western Australia (WA) is the largest single retrieval area in the world (2.6 million km^2^), with highly centralized neonatal intensive care and transport services located in the state capital of Perth – Supplement 1. The tertiary centers comprise the perinatal center at King Edward Memorial Hospital (KEMH) and the neonatal unit within the Perth Children’s Hospital (PCH). Newborn Emergency Transport Service of Western Australia (NETS WA) transfer infants from rural WA to Perth in collaboration with the Royal Flying Doctor Service (RFDS) in fixed-wing aircraft (turbo-propeller PC12 or jet PC24; Pilatus, Switzerland) Historically, non-neonatal specialist (NNS) teams transported infants to Perth due to the prohibitive travel time (slower turbo-propeller aircraft, PC12) and distance for a specialist team to transfer the infant. From 2016, following the expansion of the RFDS aeromedical fleet with jet aircraft (PC24, Pilatus, Switzerland), it was now possible for an NS (neonatal specialist) team to transport infants to Perth.

Survival of pediatric patients is improved with the use of specialized transport teams. (8) Morbidity and adverse events during the interhospital transfer of older children have long been established (9, 10). In transported pediatric patients, it has been reported that adverse events were more common when non-pediatric specialist teams performed transfer; airway events were the most common (11). There is insufficient evidence to support the positive effect of specialist neonatal teams on morbidity or mortality (12). This study aims to test the hypothesis that transports performed by neonatal specialist (NS) teams in WA have fewer unintended events than those performed by non-neonatal specialist (NNS) teams.

## Methods

All regional neonatal transports from the Kimberley and Pilbara regions of north-western Australia to Perth between 1^st^ January 2018 and 30^th^ June 2021 were identified from a prospectively collected electronic database and screened for analysis. Data were obtained from NETS WA call conference notes, clinical observation chart and patient medical records. Demographic data included gestational age, birth weight, sex, Apgar scores, and diagnoses. This study was approved through the Governance, Evidence and Knowledge Outcome oversight pathway in the Western Australia Health Department (2023-000420). Formal ethics approval was waived.

### Transport process

Neonatal retrievals included all preterm newborns (<37 weeks’ gestation) until 44 weeks corrected gestational age and term infants (≥37 weeks gestational age) until ≤28 days of age. A neonatal transport was initiated by a phone call to the specialist transport service (NETS WA) from a referring doctor, usually a pediatrician in a local hospital in the Kimberley or Pilbara, requesting neonatal transport to a tertiary facility. A call conference was convened, and clinical management was discussed. RFDS was contacted, and flights were requested. The choice of transport team was governed primarily by the infant’s location and the availability of aeromedical resources. If aeromedical resources were in the Kimberley or Pilbara region, a local pediatric doctor would accompany the infant with a RFDS flight nurse (NNS team). If a NNS team accompanied the patient from the Kimberley or Pilbara, a NS team would meet them at the airport in Perth and accompany the infant for the remainder of the journey to the tertiary unit by road ambulance (approximately 20 Km). When NNS teams accompanied the infant, a call was still required to NETS WA for clinical advice and logistical support. A neonatal transport consultant had oversight of every transport. Phone calls were documented and recorded. If the aeromedical resources were available in Perth, a NS doctor (and neonatal nurse from 2019) would travel to the patient and transfer them back to Perth (NS team).

Key time points during the transport were recorded: time of the initial referral (call to NETS WA), time of arrival of the NETS WA equipment to the cot side (in NNS transfer, the equipment would be transported from the regional RFDS base (airfield) to the cot side at the local hospital) and time of arrival in the tertiary NICU. Stabilization time was time from the arrival of the transport team and equipment to the cot side until departure from the referring hospital. Length of retrieval was the time taken from the team departure from the respective base to the infant’s arrival in NICU. The number of phone calls documented in the call conference system was recorded. Calls not documented or not made through the call conference system were not recorded or included in the data.

The principal reason for retrieval was documented. Suspected sepsis was defined based on the clinical presentation and the referring doctor’s assessment and treatment. Blood cultures and parameters confirming the diagnosis of sepsis were not available during the transport. Suspected congenital heart disease was defined as any suspected cardiac anomalies, including arrhythmias, e.g., supraventricular tachycardia. Any other congenital anomalies were defined as ‘non-cardiac congenital anomalies’.

### Physiology in transport

The Transport Risk Index of Physiologic Stability (TRIPS) score was calculated from clinical observations at the three key time points described above. The score comprises four physiological parameters (temperature, respiratory status, hemodynamic and response to noxious stimuli), reflecting neonatal physiological status at that time point. The TRIPS score predicts NICU mortality in the first seven days of life. A higher score indicates a more unstable infant with a higher predicted mortality (13).

### Team composition, definitions, and equipment

Personnel were defined and recorded as follows: Medical staff accompanying the infant were neonatologists or pediatricians. A neonatologist was a medical practitioner whose practice was entirely in that area. A pediatrician (in this context) whose practice was mainly in pediatrics but may include neonatal care. A junior practitioner was defined as less than five years and a senior more than five years of experience. A neonatal nurse had more than three years’ experience in neonatal care and postgraduate qualifications in neonatal intensive care nursing. A RFDS flight nurse/midwife (more than three years of postgraduate experience in critical or emergency care who *may* have had previous pediatric or neonatal experience) was always present on the flight (as a civil aviation requirement.) An NS team was expected to be made up of a neonatologist and neonatal nurse; an NNS team comprised a pediatrician and RFDS flight nurse.

The NETS WA equipment used to transport the neonatal patient was identical, and both teams were oriented to its use. The same equipment was stored at the regional RFDS bases in the Kimberley and Pilbara regions. Flight information was extracted from written records, including the type of aircraft (turbo-propeller or jet), maximum cruising and pressurized cabin altitudes.

### Unintended events

Unintended events during transport were documented contemporaneously, categorized as clinical, logistic or equipment and compared between NS and NNS teams. Total unintended clinical events included all events related to the endotracheal tube, hypoglycemia, hypotension, and hypo-or hyperthermia. These events were also considered separately. Hypotension was defined by the clinical need for fluid boluses or inotrope therapy as determined by the treating physician. Hypothermia was defined as <36.5°C or hyperthermia >37.5°C (axillary) on arrival in NICU. Infants undergoing therapeutic hypothermia were excluded from temperature analysis. Hypoglycemia was defined as serum glucose <47mg/dL (<2.6mmol/l) as measured with blood gas analysis. Logistic issues included delayed retrieval due to unavailability of medical, aviation or ambulance assets. Equipment issues included problems with transport cot, stretchers, and devices.

### Deaths

Deaths were reported up to seven days after admission to the receiving NICU. The reason for death was extracted from mandatory documentation and the medical notes.

### Statistics

Categorical data are presented as numbers and percentages (n, %). Comparison of categorical data is compared by Pearson’s chi-squared. Continuous variables are presented as the median and interquartile range (IQR) and compared by the Mann-Whitney U test. Comparison of the TRIPS score variables at three different time points is by a non-parametric analysis of variance with repeated measure (Friedman test). Statistical significance was considered < 0.05.

## Results

Between 1^st^ January 2018 and 30^th^ June 2021, there were 3709 emergency neonatal retrievals in Western Australia: 119 (3.2%) transports were undertaken from the Kimberley or Pilbara regions. An NS transport team transported 49 infants, and NNS transport team, 70.

### Patient characteristics

Characteristics of transported infants, reason for transfer and respiratory support on transport are described in Table 1. The NS team mainly transported infants with respiratory distress, prematurity, or a suspected congenital cardiac anomaly; NNS teams transported infants with respiratory distress or a suspected neurological or surgical disease. NS teams transported more ventilated infants than non-neonatal teams (26/49 (53%) vs. 23/70 (33%); p=0.047).

**Table 1.** Demographics of neonatal specialist teams (n=49) and non-neonatal specialist transport teams (n=70). Categorical data are presented as a number and percentage (n, %), and continuous data are presented as median (interquartile range and minimum to maximum capacity). *p=0.047

|  | NS team<br>n=49 | NNS team<br>n=70 |
| --- | --- | --- |
| Male sex | 27 (55) | 36 (51) |
| Gestation (weeks) | 38 (32 – 39; 25 – 41) | 38 (34 – 39; 25 – 41) |
| Birth weight (Kg) | 2.7 (1.9– 3.5; 0.7- 4.7) | 2.8 (2.1 – 3.2; 0.9 – 4.4) |
| Apgar Scores |  |  |
| One minute | 8 (4 – 9; 0 - 9) | 8 (3 – 9; 0 - 9) |
| Five minutes | 9 (7 – 9; 0 - 10) | 9 (6 – 9; 1 - 10) |
| Diagnosis at time of retrieval |  |  |
| Respiratory distress | 10 (20.4) | 17 (24.3) |
| Preterm | 14 (28.4) | 11 (15.7) |
| Neurology | 7 (14.3) | 11 (15.7) |
| Suspected Sepsis | 3 (6.1) | 5 (7.1) |
| Suspected Congenital Heart Disease | 8 (16.3) | 3 (4.3) |
| Non-Cardiac Congenital anomaly | 2 (4.1) | 4 (5.7) |
| Other | 1 (8.2) | 8 (11.4) |
| Respiratory support |  |  |
| Ventilation* | 26 (53.1) | 23 (32.9) |
| CPAP | 7 (14.3) | 12 (17.1) |
| HHF | 1 (2) | 1 (1.4) |
| Low flow nasal oxygen | 0 (0) | 3 (4.3) |
| No respiratory support | 15 (30.6) | 31 (44.3) |

### Physiology

Physiology scores (TRIPS) at referral, the arrival of the transport equipment at the cot side and arrival in NICU for definitive care are described in Figure 1. The TRIPS score increases across all three-time points when the infant is cared for by the NS transport team compared to the NNS team (p<0.001)—Figure 1.

**Figure 1.**
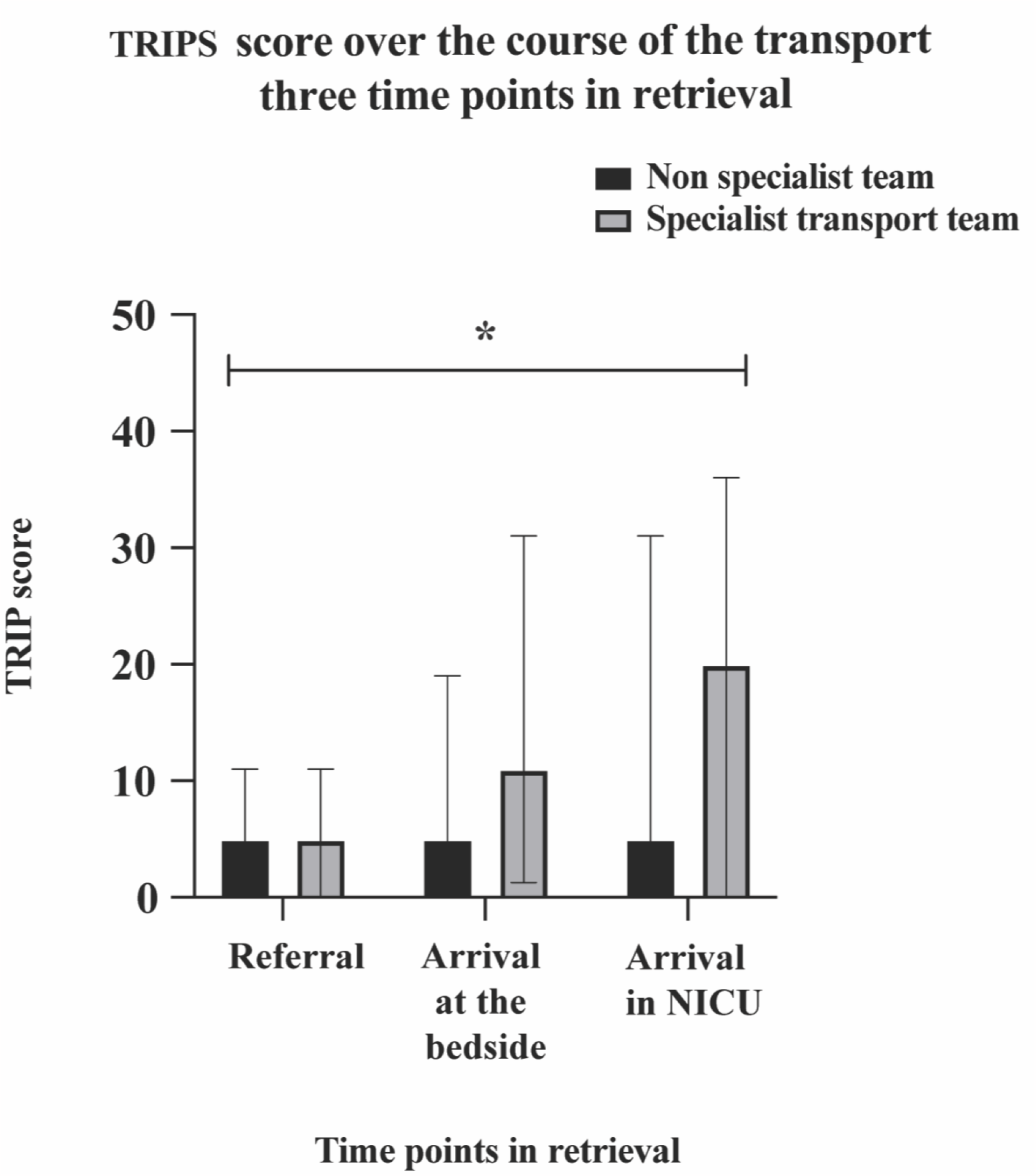
TRIPS score of non-neonatal specialist teams and specialist neonatal transport teams at the point of referral, the arrival of the NETS equipment at the cot side and the arrival of the neonate in NICU. Data are presented as median (top of solid bar) and interquartile range (whisker). * p<0.001 across the three time points.

### Transport characteristics

Transport characteristics, length of transport time and composition of teams is described in Table 2. NNS teams had shorter duration of transport times and times from decision to retrieve to arrival in NICU (508 (435 – 609) vs. 675 (615 – 735); p<0.00001 and 715 (568 – 848) vs. 871 (770 – 1030); p<0.00001) There were a greater proportion of senior medical practitioners present with NS than NNS teams (43 (88%) vs 42 (60%); p<0.001. There were more neonatal nurses present on NS (36/49 (73.5%) vs.6/70 (8.6%) p<0.001). NS teams were more likely to travel by jet than turbo-propeller aircraft (58/70 (82.9%) vs. 19/49 (39%); p<0.001).

**Table 2.**
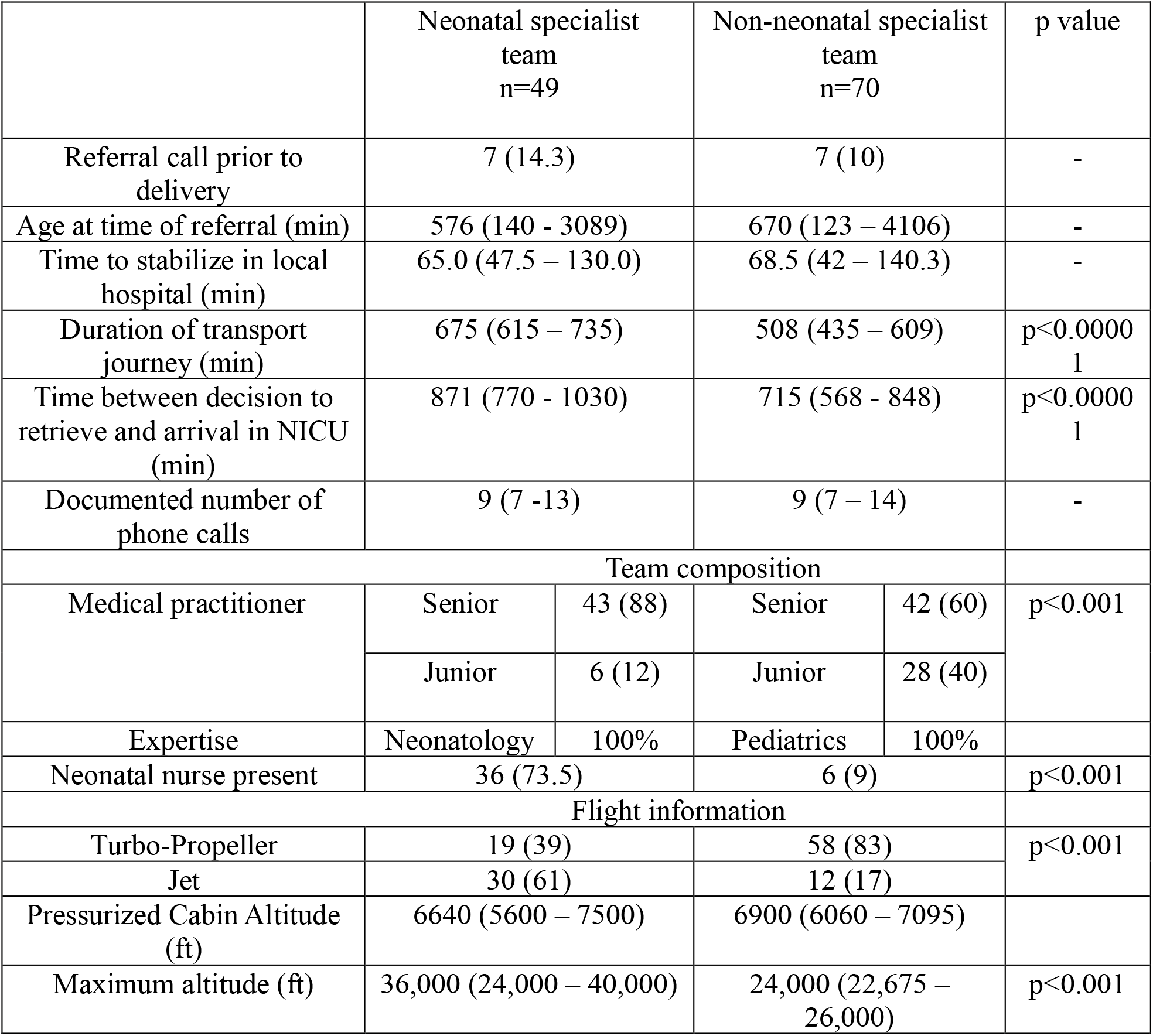
Details of transport from referral to patient arrival in NICU, the composition of the accompanying team and flight details comparing neonatal specialist transport team and non-neonatal specialist team. Categorical data are presented as numbers and (percentage) (n, %). Continuous data is presented as median and interquartile range.

### Unintended events

The total number of clinical events was less in the NS group than in the NNS group (28/49 (57%) vs. 54/70 (77%); p=0.02). There were fewer unintended events with an endotracheal tube in infants transported by NS compared to NNS teams (0/25 (0%) vs. 7/23 (30%); p=0.004). Hypoglycemia and hypo- or hyperthermia were more similar between groups. The number of logistical events was greater in the NS teams compared to the NNS group (p=0.05). Each group had a similar number of unintended equipment events. Unintended events are described in Table 3.

**Table 3.** Unintended events with hypoglycemia, temperature regulation and combined clinical, logistical and equipment events in neonatal specialist and non-neonatal specialist transport teams. All data are categorical and presented as number (n) and percentage (%).

|  | NS team<br>n=49 | NNS team<br>n=70 | p value |
| --- | --- | --- | --- |
| Hypoglycemia | 1 (2.1) | 3 (4.2) | - |
| Treated hypotension | 9 (18.4) | 10 (14.3) | - |
| Unintended temperature outside range | 12 (33.3) | 30 (47.6) | - |
| Total unintended clinical event | 28 (57) | 54 (77) | 0.02 |
| Total unintended logistical event | 31 (63) | 33(47) | 0.05 |
| Total unintended equipment event | 12 (24) | 13 (18.5) | - |

There were no deaths before the transport team arrived at the referring hospital or during the transport. Four deaths occurred within seven days of the transport, all following treatment redirection. Two-term infants with severe hypoxic-ischemic encephalopathy and two extreme preterm infants with extensive intraventricular and pulmonary hemorrhage.

## Discussion

Infants transported by NS and NNS transport teams in Western Australia between January 2018 and June 2021 demonstrated differences in team composition, unintended clinical events and logistics. NS teams transported more ventilated infants, had more neonatal nurses present during transport, and had fewer total unintended clinical events and unintended endotracheal tube events.

Few data exist to show improved outcomes in infants transported by specialist neonatal transport teams. Children transported by specialist pediatric services have fewer adverse events than non-specialist teams, especially unplanned airway events, cardiopulmonary arrest, hypotension, and loss of intravenous access (14). Consequently, pediatric specialist transport teams have demonstrated improved survival (8).

To compare neonatal transport outcomes, we selected several previously published quality metrics; dislodgement of therapeutic devices, temperature control, hypoglycemia, and hypothermia (15-18). In our study, NS and NNS teams were noted to have a difference in total unintended clinical and endotracheal tube events. We speculate that the presence of neonatal nurses as part of the transport team improved these outcomes. Neonatal nurses routinely travelled on specialist transport flights from January 2019. Before this, flight nurses were only present. This explains neonatal nurses’ <100% presence in the specialist group. No data describes the benefit of specialist nurses in transport medicine or neonatology in general. In other disciplines, it has been long recorded that specialist nurses improve outcomes (19).

The physiological condition of the infants in this study was assessed using the TRIPS score (13). TRIPS was recorded at three key time points during the retrieval. Intubated infants score highly in the TRIPS score (13), likely to be this cohort’s major contributor. The TRIPS score increased over time in the NS compared to the NNS teams. This reflects the greater potential physiological instability and medical needs of infants transported by specialist teams. The NS teams transported a higher proportion of preterm infants, those who had suspected cardiac lesions or were ventilated.

Neonatal transports from northwest WA were long journeys over many hours in all circumstances for both NS and NSS teams (675 vs. 508 min). NNS teams travelled from the referring hospital to receiving NICU. NS teams travelled double this distance. Transport times were shorter but not by half the time. NS teams were more likely to travel by jet aircraft and had more unintended logistic events than NNS. Unsurprisingly, the greater distance had more likelihood for delays due to the multiple steps in the transport journey. The longer journey time and more logistical problems accompany fewer unintended clinical events. Time-critical transports, e.g., bilious vomiting (20), require urgent and expeditious transport for definitive intervention. Patient acuity, logistics, timeliness for care and experience level of the team should be considered for the safest possible transport.

Our study is one of only a few reports comparing NS and NNS transport teams (12, 21). Strengths include reliable and comprehensive clinical status, processes, and logistics documentation from a well-maintained, prospectively acquired dataset. All transports were similar regarding equipment and consultant oversight regardless of the team composition, allowing for a like-for-like comparison. WA’s unique geographical and healthcare circumstances allow for a quasi-randomized comparison of specialist delivery in neonatal transport. Jet aircraft and the addition of a neonatal nurse to the NS team were introduced at differing time points during the study period. Group differences might have been more apparent if the specialist teams had this enhanced capability throughout the study period. Small study numbers and retrospective analysis limit our study. Due to small patient numbers, analysis of vulnerable populations and specific clinical outcomes, such as extremely preterm infants or those with hypoxic-ischemic encephalopathy, are not possible.

NNS teams in the Kimberley and Pilbara regions of Western Australia have extensive experience in providing high-quality neonatal stabilization and transport. There has long been a necessity for providing this care in WA for neonatal patients, given the limited availability of fast, long-range aircraft in WA and the distance to tertiary NICU. The acquisition of jet aircraft has provided greater opportunity for specialist neonatal teams to transport sick infants long distances in WA. Specialized neonatal nursing care appears vital in improving safety in the transport of infants.

## Data Availability

Data can be available upon request

## Abbreviations

NS: Neonatal specialist
NNS: Non-neonatal specialist
NETS WA: Newborn Emergency Transport Service of Western Australia
NICU: Neonatal Intensive Care Unit
ETT: Endotracheal tube
RFDS: Royal Flying Doctor Service of Australia
TRIPS: Transport Risk Index of Physiologic Stability
WA: Western Australia

