## Supplemental Figure 1 for "Unintended events in long-distance neonatal interhospital transport in Western Australia: A comparison of neonatal specialist and non-neonatal specialist transport teams"

Supplement 1

Figure 1. A map outlining the geographical regions of Western Australia, specifically the Pilbara and Kimberley and their relation to tertiary services centralised in Perth. The main centres for healthcare delivery are in the regional capitals of Port Hedland (Pilbara) and Broome (Kimberley). Image adapted from citation: Goulding, P M. (2014), Generalised Regions of Western Australia. Department of Primary and Regional Development, Western Australia, Perth. Map.

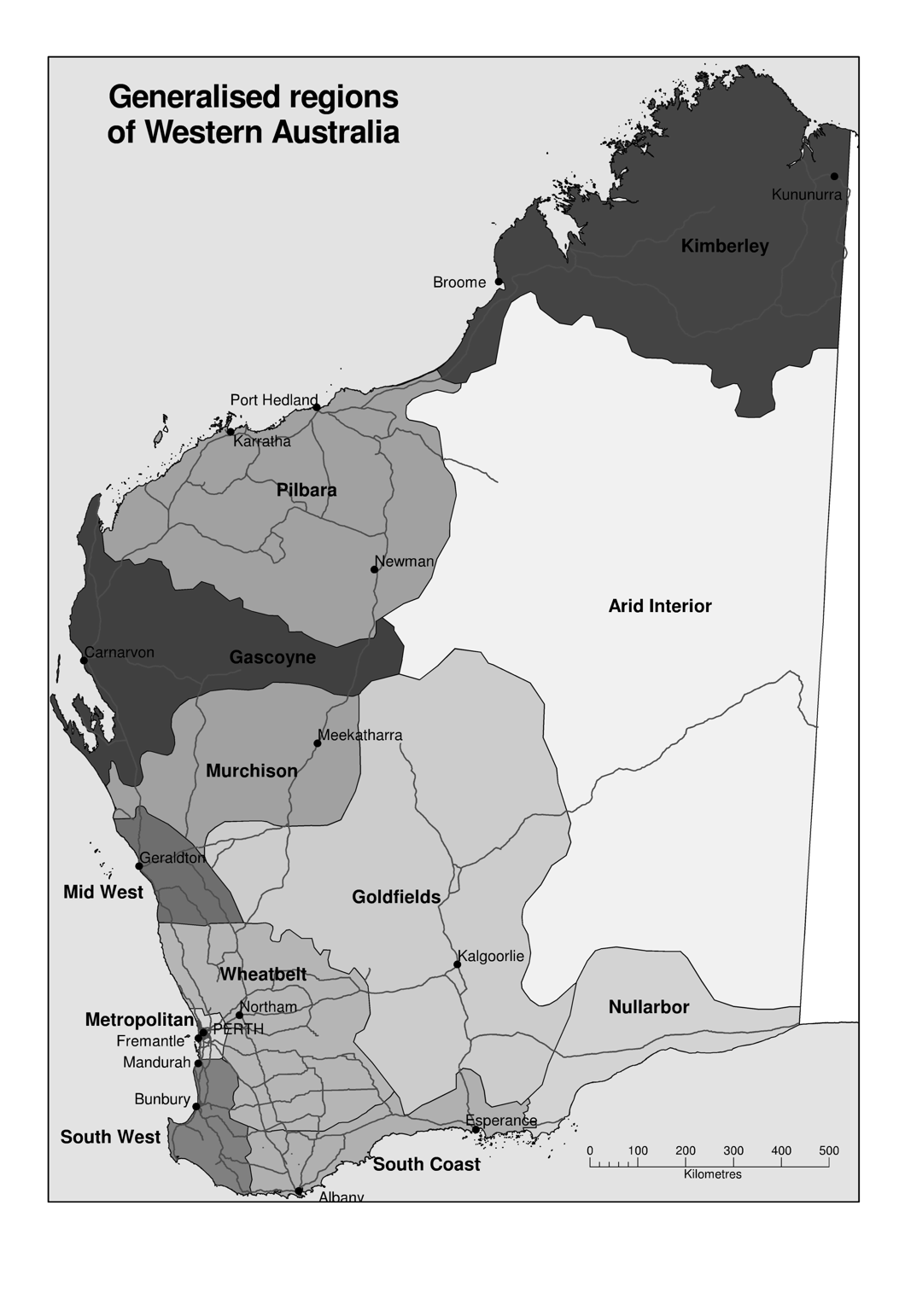
